# A novel deterministic forecast model for COVID-19 epidemic based on a single ordinary integro-differential equation

**DOI:** 10.1101/2020.04.29.20084376

**Authors:** Felix Köhler-Rieper, Claudius H. F. Röhl, Enrico De Micheli

## Abstract

In this paper we present a new approach to deterministic modelling of COVID-19 epidemic. Our model dynamics is expressed by a single prognostic variable which satisfies an integro-differential equation. All unknown parameters are described with a single, time-dependent variable *R*(*t*). We show that our model has similarities to classic compartmental models, such as SIR, and that the variable *R*(*t*) can be interpreted as a generalized effective reproduction number. The advantages of our approach are the simplicity of having only one equation, the numerical stability due to an integral formulation and the reliability since the model is formulated in terms of the most trustable statistical data variable: the number of cumulative diagnosed positive cases of COVID-19. Once this dynamic variable is calculated, other non-dynamic variables, such as the number of heavy cases (hospital beds), the number of intensive-care cases (ICUs) and the fatalities, can be derived from it using a similarly stable, integral approach. The formulation with a single equation allows us to calculate from real data the values of the sample effective reproduction number, which can then be fitted. Extrapolated values of *R*(*t*) can be used in the model to make reliable forecasts, though under the assumption that measures for reducing infections are maintained. We have applied our model to more than 15 countries and the ongoing results are available on a web-based platform [1]. In this paper, we focus on the data for two exemplary countries, Italy and Germany, and show that the model is capable of reproducing the course of the epidemic in the past and forecasting its course for a period of four to five weeks with a reasonable numerical stability.

## 1 Introduction

Our primary aim is to set up a deterministic model that can be easily tuned with available data in order to make numerically stable forecasts. We found that existing methods are not well-suited to reach this goal.

Empirical top-down modelling, *i*.*e*., approaches that start from data and make prognoses, mostly ignore underlying dynamics. The easiest approach is curve fitting of available data. In [2] the number of cumulative diagnosed positive COVID-19 cases *P* (*t*) was assumed to be an error function. This is true if the number of daily new cases *P′* (*t*) (prime denotes derivative with respect to *t*) can be described by a Gaussian distribution. As we will show, a symmetric distribution function *P′* (*t*) corresponds to an effective reproduction number that converges rapidly to zero. This might be true for China data. In Italy and Germany we observed a final (at the time of writing) value for *R*_eff_ between 0.6 to 0.8, leading to an asymmetric function *P′* (*t*) with a long tail. Although the peak date has been predicted well in [2], the predicted total cases and fatalities differ by more than 30%.

Current deterministic models were developed with the aim of simulating possible scenarios and showing the effect of containment and mitigation measurements. They are “bottom-up” in the sense that they are based on the knowledge of typical epidemiological parameters, such as the basic reproduction number *R*_0_ or the time between contacts *T*_*c*_, just to name a few. However, three reasons make it difficult to set up these complex models for forecasting:

− the epidemiological parameters are unknown and change in time;
− for most of the compartmental model variables, such as susceptible, exposed, infected or removed individuals, the availability of surveillance data is limited;
− model tuning requires fitting many variables simultaneously - making it difficult to find an optimum.

In [3] the classical SIR model has been applied to Italy, dividing the country into three parts: north, centre and south. The problem they face, in our opinion, is that the official number of infected individuals *I* contains people who are officially not cured. But in Italy people enter the statistics as cured when they have been tested as negative twice or even three times in a week’s distance. Thus, from a dynamic point of view they remain “infected” for too long. The model cannot capture this feature appropriately and, in order to keep track of the statistical data, it has to be re-tuned within days.

The German Robert Koch-Institut (RKI) uses an extended SEIR model to show various scenarios for the course of the COVID-19 epidemic in Germany by applying different seasonality of the epidemic and immunity of the population [4].

Another Italian team has set up a model with eight prognostic variables, SIDARTHE [5], taking also into account asymptomatic cases and detection issues. Again, these efforts allow precise simulation of scenarios but are difficult to be set up with real data to make forecasts. The comparison with real data looks good but is restricted to the initial period of the epidemic when the case numbers grew simply exponential.

In [6] statistical parameters are obtained to feed parametric models, though not explicitly specified. Ensemble calculations using various data sources and different models allow for evaluating the statistical spread of the obtained forecasts - a procedure which is widely used in meteorological forecasting. The overall approach seems successful but remains complex.

Estimates of the disease transmissibility obtained through the evaluation of the time-dependent *reproduction number R*_*t*_ have been proposed by various authors (see, e.g., [7–9]. Wallinga and Teunis [7] proposed a statistical approach to compute an effective reproduction number which requires as input only the number of daily cases and the distribution of times intervals between the appearance of symptoms in primary cases and the onset of symptoms of secondary cases. The main drawback of this method is that in order to obtain estimates of *R* at time *t*, incidence data from times later than *t* are required.

To check the efficacy of restrictive measures adopted to contrast infectious disease, Bayesian estimation of the reproduction number *R*_*t*_ along with Markov chain Monte Carlo and Monte Carlo sampling are employed in [8] to infer the temporal pattern of *R*_*t*_ up to the last observation.

Cori et al. define in [9] the instantaneous reproduction number *R*_*t*_ as the ratio of the new infections at time *t* to the total infectiousness of infected individuals at the same time. In this way, *R*_*t*_ represents the average number of secondary cases that each infected individual would infect if the conditions remained as they were at time *t*. It is interesting to note that infectiousness of each individual is modulated by a weighting infectivity function which mainly depends on individual biological factors such as pathogen shedding (see also next eq. (4)). Forecasting is then based on Bayesian statistical inference which leads to a simple expression of the posterior distribution of *R*_*t*_, assuming a prior gamma distribution for *R*_*t*_ [9].

If data-based forecasts are the primary scope, it seemed reasonable to us developing a hybrid approach: a simple dynamic model that can be easily tuned with available data. This goal is obtained with our approach based on a single prognostic variable, which is found to satisfy an ordinary integro-differential equation.

To our knowledge, there are only a few approaches that are equally simple and effective. In [10] a delay model is presented with a single prognostic equation that has even an analytic solution. Arguments and results are comparable to ours, though our integral formulation is more general and more robust when extracting parameters from available data to feed the prognostic model. Delay models [11, 12] can also be described with several variables and many parameters, which again makes them difficult to set up as forecasting model.

The paper is organized as follows. In Sect. 2 we derive the model and show that it can be interpreted as a generalisation of classical compartmental models, such as SIR. Section 3 is devoted to the analysis of real data from the COVID-19 epidemic in Italy and Germany. A summary of how the model is capable of handling data from other countries is given in Sect. 4. We conclude the presentation of our model with some remarks on stability and numerical robustness in Sect. 5. Finally, some conclusion are drawn in Sect. 6.

## 2 The model

### 2.1 Derivation of model equations

Many compartmental models, such as SIR, use deterministic equations for susceptible *S*, removed *R* and currently infected individuals *I* - all these variables being difficult to obtain from real data for various reasons. In our opinion, the most reliable statistic variable is the *number of cumulative diagnosed positive cases*. We choose this quantity as our model variable and denote it by *P*. We are aware of the fact that the diagnosed cases are only a part of all positive cases but we assume that:

i. they are a statistically relevant part of the population;
ii. the fraction of diagnosed to all cases does not change through time and therefore,
iii. the dynamics applied to the *visible* part of the epidemic is representative for the entire epidemic.

Concerning point (ii), at the early stages of the epidemic the fraction of diagnosed cases obviously increased, also in response to the rapid increasing of the number of tests, reaching then an approximate stationary value. Variations of this stationary value are however negligeable since it soon appeared that a large percentage (up to 80%, depending on the area) of people testing positive for COVID-19 may be asymptomatic. Therefore, symptom-based screenings, which are the most frequent epidemiologic investigations adopted in many countries, are likely to miss a lot of them (see [13] and references therein). Though the diagnosed cases are only a fraction of the total cases, what is important is that the proportion of asymptomatic cases be nearly constant over time (in this regard, see also the interesting simulation study reported in the Web Appendix 8 of [9]).

One of the main objectives of an epidemiological analysis is to give estimates of the reproduction number *R* after restrictive measures (e.g., confinement, social distancing) have been adopted to limit epidemic spreading. With such constraints, assuming constant environment and exponential increase in new case counts appears unjustified [14] and a more empirical data-based approach is more appropriate to follow the temporal evolution of the reproducing number. We derive our model in a discrete version, using discrete daily values as they are given by various data sources. Successively, in Sect. 2.2 we illustrate its continuous version.

We refer to *P*_*n*_ as the number of cumulative diagnosed positive cases on day *n* and to *ΔP*_*n*_ as the number of newly infected COVID-19 cases on day *n*. We denote by 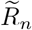 the ratio between the new cases *ΔP*_*n*_ on day *n* and the weighted sum of new cases on the previous *N*_*r*_ days:

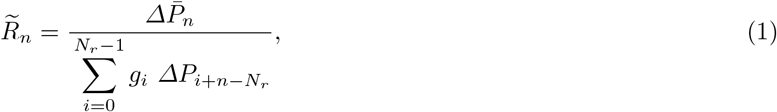

{*g*_*i*_} being a set of *N*_*r*_ fixed weights with the property 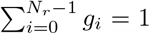, where *N*_*r*_ is the average number of days until an infectious person is removed from the infection process and 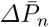 is a suitable average of *ΔP*_*n*_.

Let us first illustrate the main idea supporting our hybrid approach. The numbers 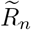 can be calculated easily from the existing epidemic data. Then, a regression curve *R*(*t*) can be fitted to the set of numbers 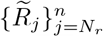, thus providing us with a law, purely based on data, on how this epidemic variable evolves. The data we observed show that, prior that restrictive measures have been adopted,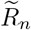 has a nearly constant initial value corresponding to the basic reproduction number *R*_0_. Once contact behaviour changes (due to media information, measurements, quarantine, and so on) from a certain time *T*_Q_ on, 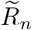 is no longer constant but manifests an evident decay towards a final asymptotic value. Therefore we are prompted to choose the following model to describe the data 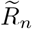:

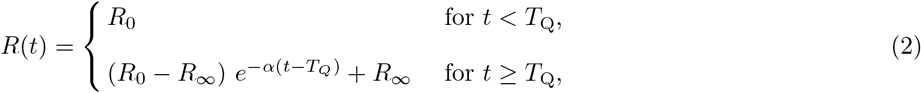

*α, R*_0_ and *R*_*∞*_ being parameters to be determined from data 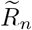. Note the difference between the discrete values 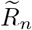, calculated from data by eq. (1), which show sample fluctuations and the numbers *R*_*n*_ *≡ R*(*n*), which are the values of the regression curve evaluated at the sample day *t ≡ n*. Also note that phases of different severity in mitigation measures lead to small intermediate plateaus in the time course for 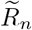. This behaviour of the data was also noted in [15], especially for what concerns fatalities. Since the steps in 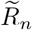 are strongly smeared out, we simply model data by one single step with an asymptotic decay. Obviously, in order to gain higher accuracy, *R*(*t*) could be described with a more complicated, piece-wise defined function but at the price of introducing additional fitting parameters. The time *T*_*Q*_ can be read rather easily from the course of the data and set approximately as the time when 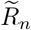 starts decreasing. Moreover, we will see in the next section that in the special case of constant weights *g*_*i*_ = 1*/N*_*r*_, the quantity *R*(*t*) is actually seen to have the meaning of an effective reproduction number.

Resolved for 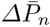, eq. (1) turns into a prognostic model for future infection cases *ΔP*_*n*_ but provides also a model-based, *smoothed* curve for representing present and past data, *i*.*e*.:

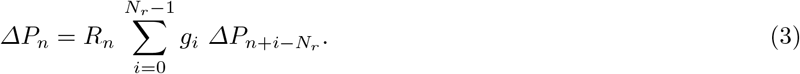

Introducing weights *g*_*i*_ is *sensible* because clinical data show that the probability to infect others is not equally distributed over time. The incubation time is known to be between 1 and 14 days, with an average of 5 days [4]. The infectiousness begins probably before symptoms manifest and is maximal at the beginning of the disease. All these characteristics can be captured with a suitable choice of the summation weights *g*_*i*_.

The weights *g*_*i*_ are samples of an infectivity probability (see, e.g., Fig. 2) over the time period of *N*_*r*_ days in the past (*i*.*e*., *t ≤* 0) that we have assumed to be Gamma-distributed with shape *p* and rate *b*. The corresponding probability density function then reads:

**Fig 1:**
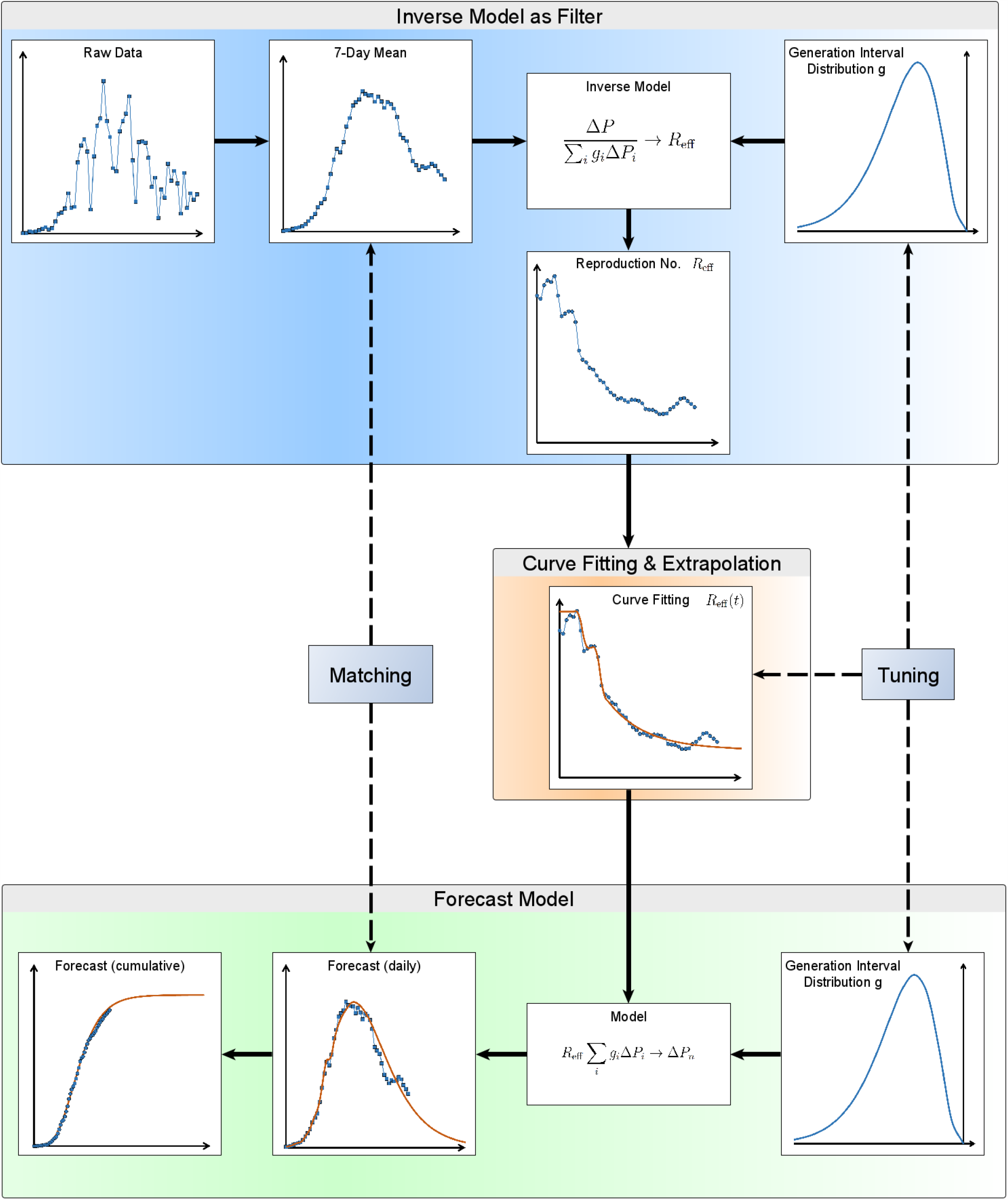
*Flow chart of the proposed model*.

**Fig 2:**
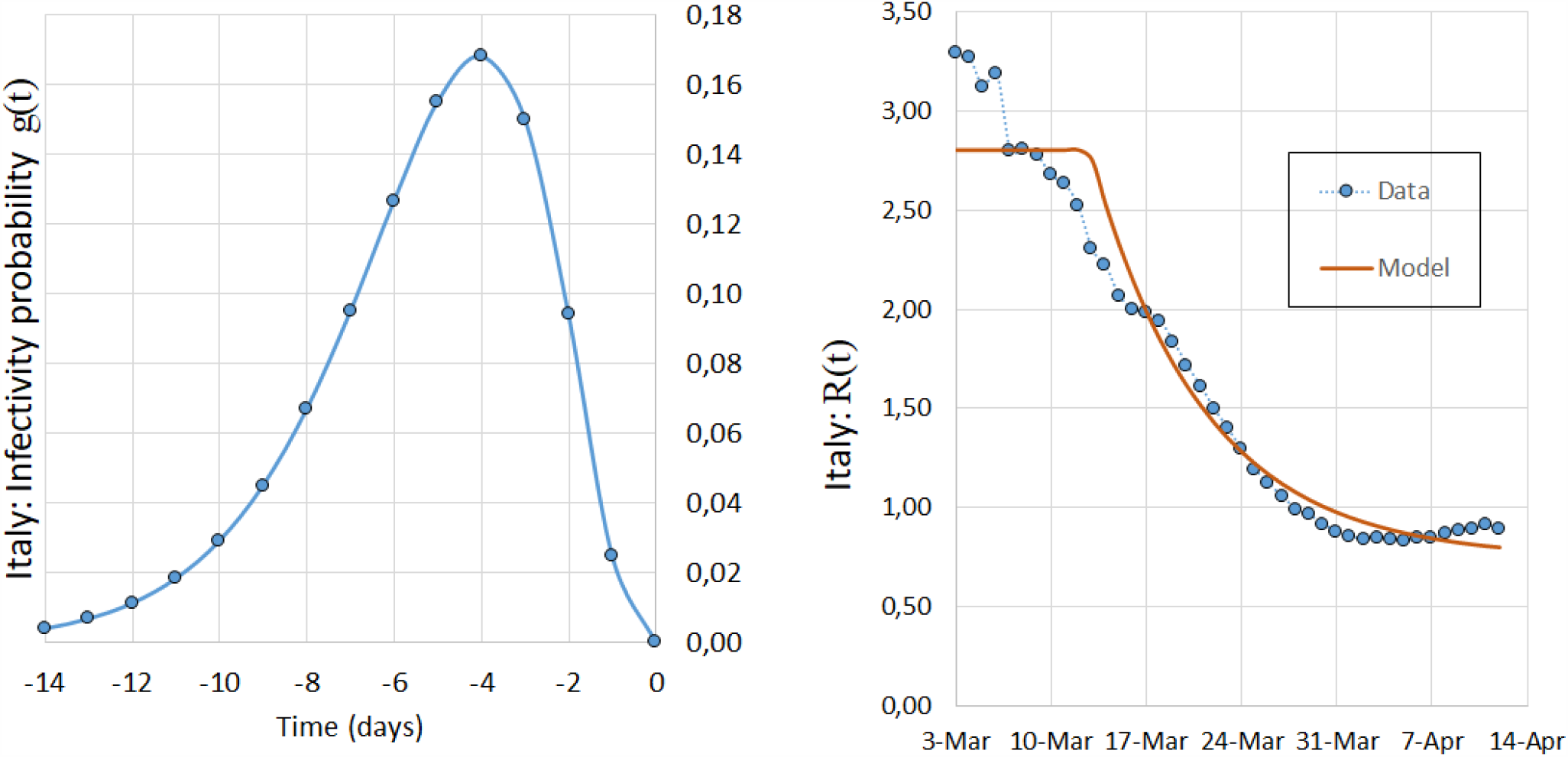
*Left: Γ -distributed weights used to calculate* 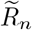 *with p* = 4 *and b* = 0.75 *(see eq*. (4)*). Right: Effective reproduction number R*(*t*) *obtained from data* 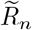 *(blue line) and the fitted curve (see eq*. (2)*) (orange line) used to model the epidemic in Italy. R*_0_ = 2.80, *α* = 0.12, *R*_*∞*_ = 0.75.

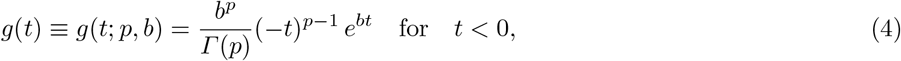

*b* and *p* being parameters to be fixed on the basis of the biological/clinical data of the disease. Note the definition of *g*(*t*) for negative values of *t* in order to provide probability values for the “past time” (see next eqs. (5) and (6)). A Gamma distribution with suitable parameters describes well what we know about the temporal distribution of infectiousness of the disease: no or low infectiousness in the first few days, a rapid slope towards a maximum followed by a slow decay. Assuming that the infectiousness of a single individual is Gamma-distributed, the infectiousness of the sum of all individuals is again Gamma-distributed and therefore also the average infectiousness used in our deterministic model has this characteristic distribution. The Gamma distribution in the context of epidemic modelling was introduced in [16] for stochastic epidemic models and later applied in [17] for the derivation of quasi-stationary distributions of the SIS and SEIS model. The values of the parameters *N*_*r*_, *b* and *p* are given by the clinical observations and are typical of the disease. In Sect. 3, where we discuss the epidemiologic data, we used *N*_*r*_ = 14, *b* = 0.75 and *p* = 4, which corresponds to the peak of infectivity after 4 days.

Therefore, we have introduced a prognostic model with six degrees of freedom: three degree of freedom set by the clinical information, *i*.*e*., *N*_*r*_ representing the infection or removal time in days, *p* and *b* for describing the infectivity probability; the other three degrees of freedom *R*_0_, *α* and *R*_*∞*_, obtained by fitting the regression curve *R*(*t*) in (2) to the data 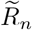, represent the time dependance of the effective reproduction number upon the social restrictive measures. The logical steps associated with the proposed model are summarized in Fig. 1.

The fitting parameters entering the model can be seen either as pure tuning numbers and also can be interpreted in epidemiological terms. In fact, the model as a whole can be compared to standard compartmental models, such as SIR, as we will show in the next section.

### 2.2 Comparison to other models

In order to compare our discrete model to classical continuous deterministic models, we write eq. (3) in the following continuous form:

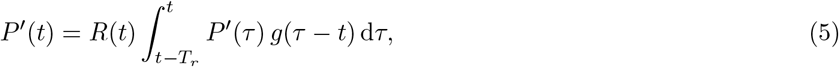

where *P* (*t*) is the number of total COVID-19 cases, prime stands for derivative, *R*(*t*) represents, as we will show, an effective reproduction number, *T*_*r*_ is the continuous generalization of *N*_*r*_ given in eq. (1) and represents the time during which infected individuals take part in the infection process, and *g*(*t*) is a weighting function representing the infectivity probability:

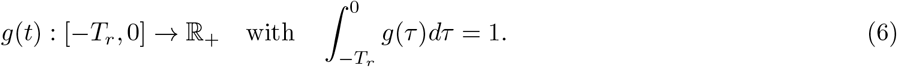

Notice that *g*(*t*) is defined for negative values of *t*, emphasizing thus the fact that the averaging process described by the integral in (5) works over past times or, roughly speaking, that an individual found infected at time *t* (secondary case) has been actually infected some time before. Finally, recall that the numbers *g*_*i*_, which appear in (1) and (3), are samples of probability distribution *g*(*t*). It is worth observing that eq. (5), read as the equation ruling *R*(*t*):

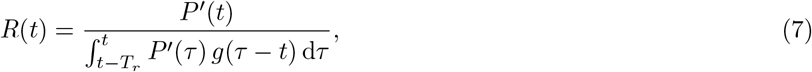

can be obtained as a particular case of the renewal equation for the birth process, where *P* ^*t*^(*t*) represents the observed birth rates and the time-varying infection rate *R*(*t*)*g*(*− τ*) refers to the rate of production by a mother at the (positive) age *τ* [14, 18].

### The SIR model

The SIR model, initiated by Kermack and McKendrick in 1927 to describe the plague spread mechanisms in Mumbai [19], is a classic compartmental epidemic model that works with three prognostic time-dependent variables: the susceptible individuals *S*(*t*), the infected individuals *I*(*t*) and the people removed from the infection process *r*(*t*). There are transitions from *S* to *I* to *r*, which lead to the following system of ODE’s [20]:

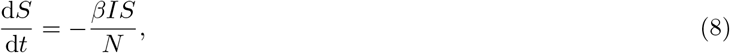

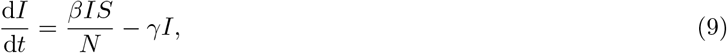

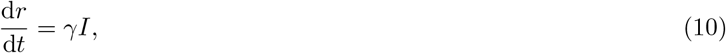

where *N* = *S* + *I* + *r* is the total number of population, *β* = 1*/T*_*c*_ is the contact frequency, *T*_*c*_ being the average time between contacts and 1*/γ* = *T*_*r*_ is the mean time between infection and removal. We skip here the discussion of the SIR model (for further details, the interested reader is referred to [21]) and ask: How does our model compare to SIR model? For this purpose, let us rewrite the SIR model in terms of our prognostic variable *P* (*t*) = *I*(*t*) + *r*(*t*). If we add Eqs. (9) and (10) we obtain

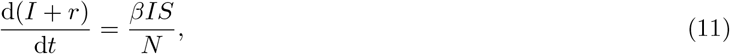

which, in terms of *P* (*t*), becomes

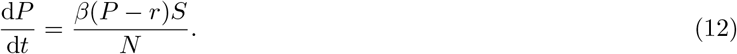

Introducing relative susceptible number *s* = *S/N*, eq. (12) can be written in the following form:

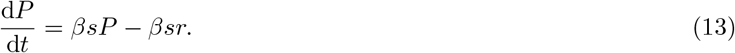

Now, let us rewrite our model (see eq. (5)) by splitting the integral into two parts and replacing *g*(*t*) with constant weights *g*_0_ = 1*/T*_*r*_. We have:

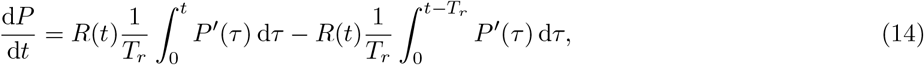

which, after integration, yields

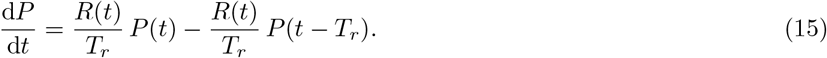

Comparing the SIR model in the form (13) with our model (15), we can identify

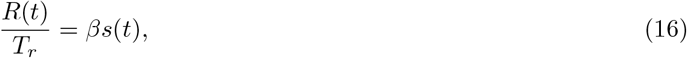

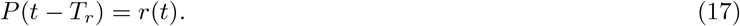

Recalling the standard definitions, *γ* = 1*/T*_*r*_, *β* = 1*/T*_*c*_ and introducing the basic reproduction number *R*_0_ = *T*_*r*_*/T*_*c*_ = *β/γ*, we can state the following epidemiological interpretation of the parameters:

i. Since 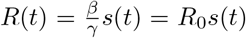, the quantity *R*(*t*) assumes indeed the meaning of a time-dependent effective reproduction number: *R*(*t*) *≡ R*_eff_ (*t*) = *R*_0_*s*(*t*).
ii. The positive cases *P* (*t− T*_*r*_) correspond to the removed individuals *r*(*t*), thus *T*_*r*_ can be consequently interpreted as the time until removal from the infection process.

In general, *g*(*t*) is not constant and it is typical of the disease. In this case, we can extend what discussed in point (i) above and give to *R*(*t*) the meaning of *generalized effective reproduction number* associated with the probability distribution *g*(*t*).

### Delay models

Delay models [22, 23] follow essentially the same strategy as SIR, the main difference being that the removal process is not modelled by a separate variable but with a time shift in the function describing the number of cumulative cases. In fact, eq. (15) is identical to the functional retarded differential equation (11) in [10], with the averaging weights *g*_*i*_ set to the constant value *g*_*i*_ = 1*/T*_*r*_.

### The SEIR models

The SEIR models (see [20] for a comprehensive review of the fundamental dynamics of these models) introduce a further group of people, the *exposed* E, that is, people who are infected but not yet infectious. This effect is accounted for in our model by excluding the first days into the integral in eq. (5) or, equivalently, by using null weights, *g*_*i*_ = 0, for those days. Though, from the clinical observation of the COVID-19 epidemic it seems that the probability that people are infectious already from the first days after infection [4].

### What makes the difference?

1. First of all, our approach does not explicitly model *S*(*t*) with a coupled prognostic equation. This sounds reasonable to us because the assumption that susceptible individuals are removed only by the infection process is wrong for the current COVID-19 epidemic. In fact, severe quarantine measures, including lockdowns and social distancing, have been implemented in almost all countries.
2. Other compartmental models take such measures into account by introducing, e.g., direct transfers from *S* to *r* compartments. However, in our opinion, this makes these models complicated and hides the fact that political measures and their effects are extremely difficult to model. Our approach is a very practical one: we do not model *S*(*t*). We focus on *R*(*t*), which we have seen being related to the product of the basic reproduction number *R*_0_ and the time-dependent relative susceptible number *s*(*t*). We extract *R*(*t*) from real data using our model assumptions and apply a curve fitting procedure to allow for extrapolation. Therefore, our approach can be called “hybrid”: a mixture of curve fitting and modelling.
3. By using the number of cumulative diagnosed positive cases *P* (*t*) as prognostic variable, we automatically have the numbers of new infections as *ΔP*_*n*_. In our opinion, this is the best variable to describe how the epidemic evolves. In SIR models, this number is not automatically obtained since *I* represents the “currently infected people” and *ΔI* is a net difference mixing the “new positive cases”, that is, the transfer from *S* to *I*, with the “removed cases”, *i*.*e*., the transfer from *I* to *r*.
4. By introducing the weights *g*_*i*_ we can model the incubation time, namely, the time between “getting infected” and “being infectious”, as well as the time before detection.
5. We define the removed people *r*(*t*), which counts the individuals that no longer take part in the infection process, as all positive cases *P* (*t − T*_*r*_) at a certain previous time *T*_*r*_. Again, we have no prognostic equation for *r*(*t*). Therefore, we do not have to model and elaborate how these individuals are removed from the infection process. We simply assume they are removed after a certain time *T*_*r*_ because, prior to curing or deceasing, people are isolated in hospitals or, in the case of weak symptoms, are put into quarantine.
6. Parameters or variables that are not known, such as *T*_*c*_, *R*_0_, *S*(0) and *N*, are subsumed into a single function *R*(*t*), which is obtained from real data, without having to speculate on how it comes about. This makes the model simple - and setting the weights constant - even simpler so that it can be set up quickly to produce satisfying prognoses.
7. From the comparison with the compartmental SIR models, we see that our epidemiological variable *R*(*t*) assume the meaning of *generalized effective reproduction number* associated with the infectivity probability *g*(*t*), which is typical of the disease.

With these assumptions we have been able to describe the infection process with a single prognostic variable *P* (*t*) in an integro-differential equation. From our perspective, the computation of deceased and cured people is a secondary process, which does not influence the dynamics of the epidemic. Nonetheless, they are important numbers to know and, however, they can be simply obtained from *P* (*t*), as it will be shown in the next section.

### 2.3 Secondary variables

The analysis of the “number of fatalities”, “number of cured” and “number of active cases” follows *mutatis mutandis* the analysis presented in Sect. 2.

### The number of fatalities

Let *V*_*n*_ be the total number of deceased individuals on day *n* from the beginning of the epidemic. We assume that the casualties on the *n*th day, *i*.*e. ΔV*_*n*_, is related to the weighted sum of new cases over the last *N*_*V*_ days, yielding the definition of the following empirical ratio:

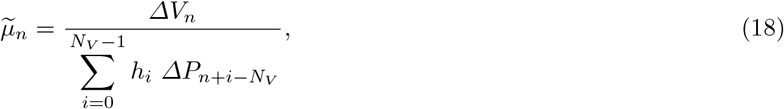

where *N*_*V*_ is the maximum number of days after which the people decease and the weights *h*_*i*_ allow for taking into account a probability distribution of deceasing. The numbers 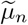 can be interpreted as case fatality ratios [24]. Similarly to what we have done in the Sect. 2, the values 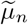 can be computed from existing data, and then fitted to a model function *µ*(*t*) to extrapolate future values: *µ*_*n*_ *≡ µ*(*t*_*n*_), where *t*_*n*_ is a day in the future. The corresponding discrete prognostic equation reads

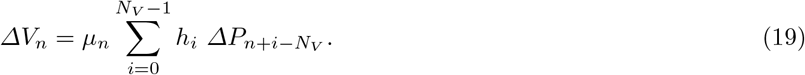

Note that from the analysis of currently available data, it results that the peak of fatalities lacks about 7-8 days behind the peak of the daily new infections. This has been accounted for in the weights *h*_*i*_, having set a maximum value at 7-8 days prior to the current day *t*_*n*_.

### The Number of cured

Let *C*_*n*_ denote the number of currently cured individual and *ΔC*_*n*_ the number of new cured individuals at day *n*. The empirical discrete curing ratio *ν*-_*n*_ can be defined analogously (see Eqs. 1 and 18):

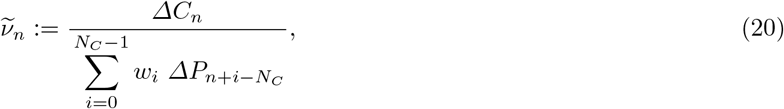

where, as in the previous cases, *w*_*i*_ denote suitable weights. Even in this case, the discrete curing ratio can be fitted with a suitable model function *ν*(*t*) to obtain predictions: *ν*_*n*_ *≡ ν*(*t*_*n*_), which can be used to predict future number of cured people through the relation

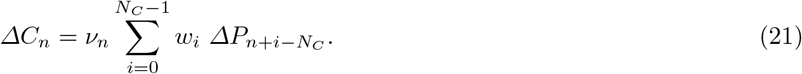

### Active Cases

The number of currently infected individuals *I*(*t*), also known as active cases, is the difference between all cases *P* (*t*) and the deceased and cured cases: *I*(*t*) = *P* (*t*) *− C*(*t*) *−V* (*t*). Note that calculating the number of removed cases in the usual way is not valid for our model, that is, we have:

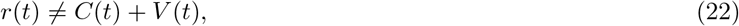

since our removal process is obtained by cutting off the corresponding integral after a removal time *T*_*r*_, taking thus into account not only the usual removal processes due to curing and deceasing, but including even other processes such as quarantine or isolation. However, we have to guarantee by means of the suitable choice of our tuning parameters that, in the long term,

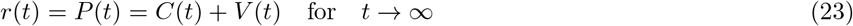

since, in this limit, *I*(*t*) = 0.

## 3 Discussion of model results

Data for the COVID-19 epidemic are made available by the John Hopkins University [25] and coincide, at least for Italy and Germany, with those from Worldometers [26]. The original time series data show significant weekly fluctuations, hence we only work with 7-day averages, which acts as a low-pass filter.

### 3.1 Italy

On the left of Fig. 2 we see the *Γ* −distributed weights *g*_*i*_ we used to obtain the effective reproduction numbers 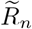 (see eq. (1) and the figure caption for numerical details) shown on the right of the same figure. The integration time is *N*_*r*_ = 14 days, *i*.*e*. only individuals registered positive within this time period actually take part in the (model) infection process.

The effective reproduction numbers 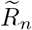 are shown for the time period from March, 3 to April, 14. The data-based 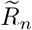 are shown with blue dots and the model-fitting curve with the orange line. The data shows two short plateaus. The first one at *R*_0_ *≈* 3.3 represents the basic reproduction number before any restrictive measures. The second plateau at *R*_0_ *≈* 2.8 represents an intermediate reproduction number before lock down. After three weeks the data 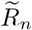 seem to settle at a value of about 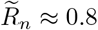. For simplicity we fitted this behaviour with only one step by eq. (2), the resulting parameters being: *R*_0_ = 2.80, *α* = 0.12, *R*_*∞*_ = 0.75.

#### The COVID-19 cases in Italy

The daily new diagnosed cases are shown on the left of Fig. 3. The original time series data show significant weekly fluctuations, therefore we show only the 7-day average. The model is capable of reproducing the exponential growth in the beginning of the epidemic, as well as the peak and the slow decay of the curve afterwards. The deviation of the curve *ΔP* (*t*) from the data 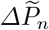 is the consequence of the deviation between 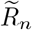 and fitting curve *R*(*t*). The cumulative number of diagnosed positive cases *P* (*t*) are shown on the right of Fig. 3. The model curve *P* (*t*) follows rather accurately the data 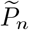. Note that the last model-tuning was made on April 13, 2020. Two weeks later the relative deviation of the cumulative number of cases is about 2%. Approximately 2.5 months later, at the moment of revision, using the tuning of April, 13 we had a deviation of less than 10%.

**Fig 3:**
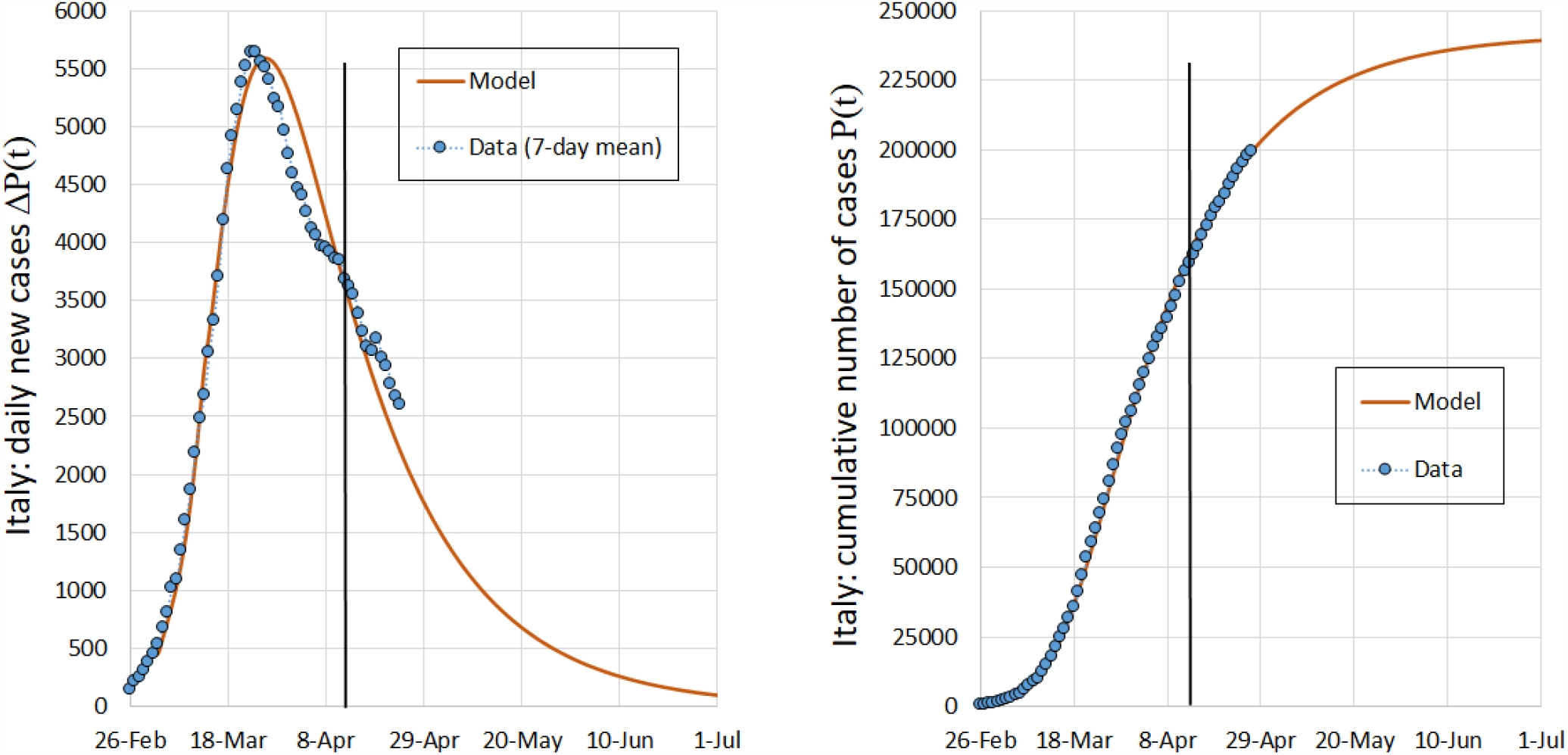
*Daily positive diagnosed cases ΔP* (*t*) *(left) and number of cumulative positive diagnosed cases P* (*t*) *(right) from February 26 to July 1, 2020 in Italy. Only data before the vertical black line, i*.*e*., *before April 13, have been used to tune the model*.

#### Fatalities in Italy

On the left of Fig. 4 we see the Gaussian weights *h*_*i*_ (see eq. (18)) used to obtain the model case fatality rate 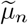 shown on the right of the same figure. The integration time is *N*_*r*_ = 18 days, *i*.*e*., only individuals diagnosed positive within the last 18 days are considered in the model calculation of fatalities. Note that setting the Gaussian time-shift, that is the location of the Gaussian peak, at *t*_shift_ = 6 puts the maximum weight on patients that have been diagnosed positive 6 days before. This is sensible because the peak of daily fatalities occurs 6 days after the peak of daily new infections. The number of daily *ΔV* (*t*) and cumulative fatalities *V* (*t*) are shown in Fig. 5. The course of fatalities is well represented by the model and 14 days after the last model-tuning the relative deviation for the cumulative number of fatalities is about 2%.

**Fig 4:**
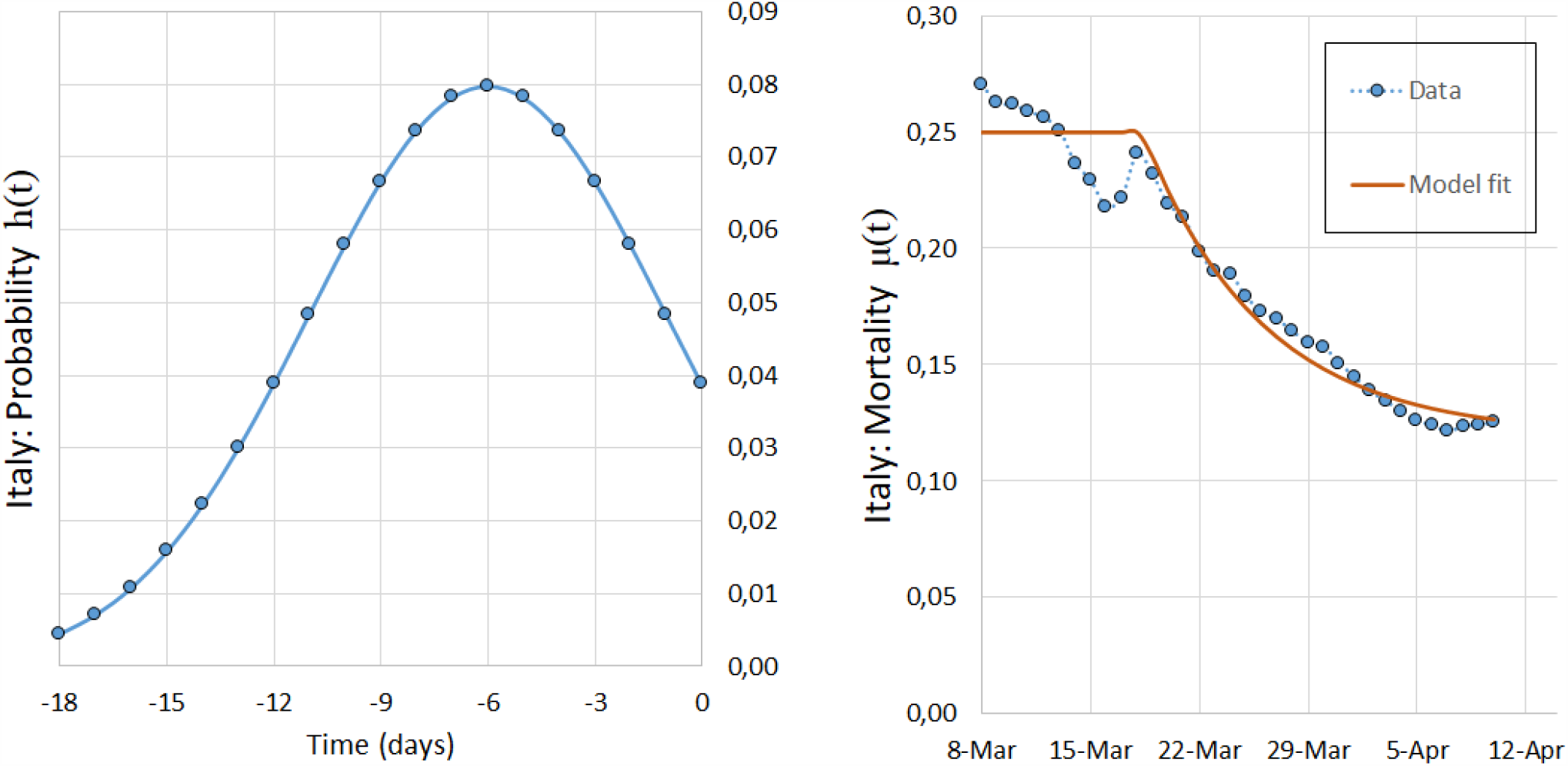
*Left: Gaussian weights h*_*i*_ *used to calculate* 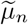 *with σ* = 5 *and t*_shift_ = 6. *Right: case fatality rate µ*(*t*) *obtained from data* 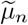 *(blue points) and the fitted curve (orange line) used to model the epidemic in Italy*.

**Fig 5:**
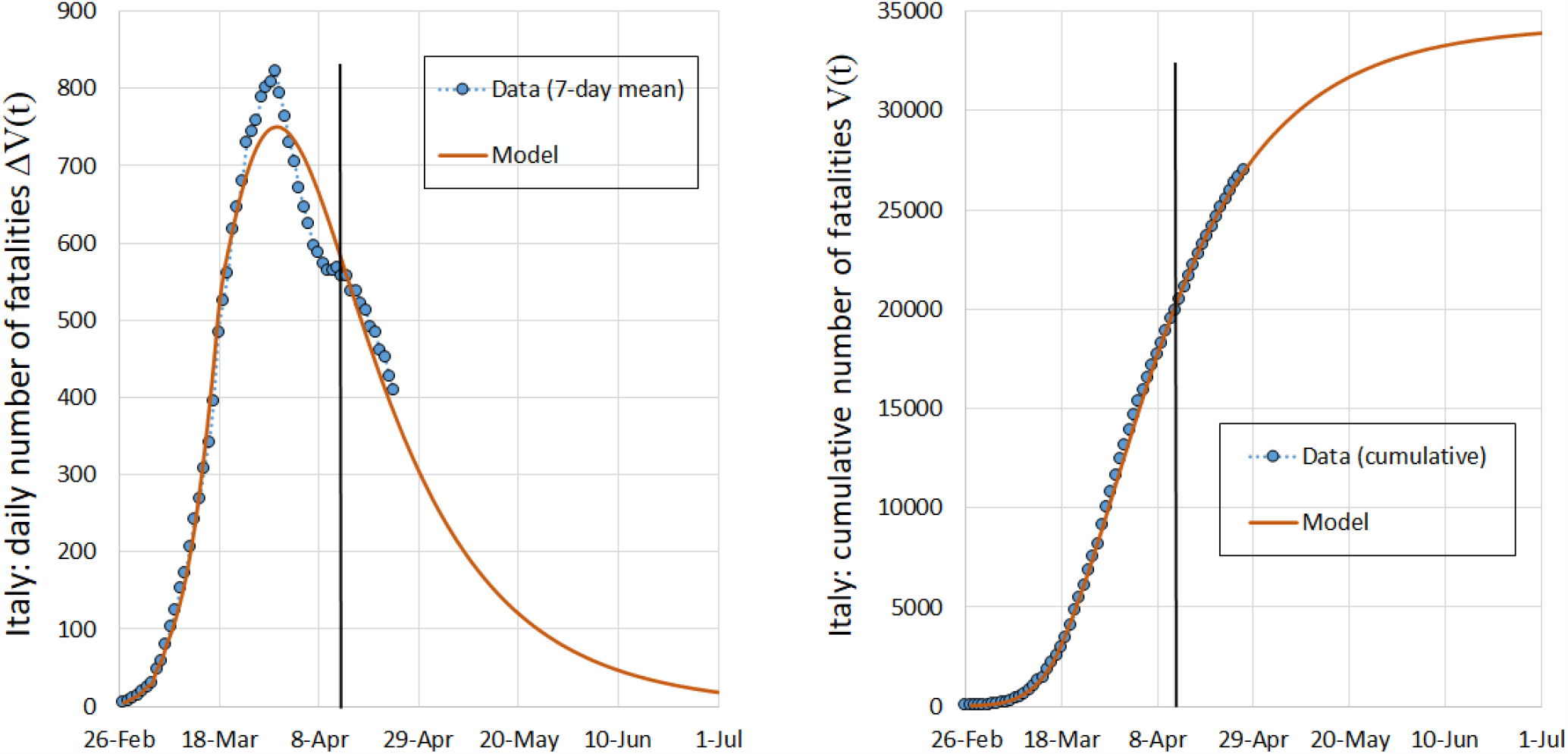
*Daily new fatalities ΔV* (*t*) *(left) and cumulative number of fatalities V* (*t*) *(right) from February 26 to July 1, 2020 in Italy. Only data before the vertical black line, i*.*e*., *before April 13, have been used to tune the model*.

### 3.2 Germany

On the left of Fig. 6 we see the *Γ −*distributed weights *g*_*i*_ we used to obtain the effective reproduction numbers 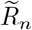 shown on the right panel. Again, the integration time is *N*_*r*_ = 11 days.

**Fig 6:**
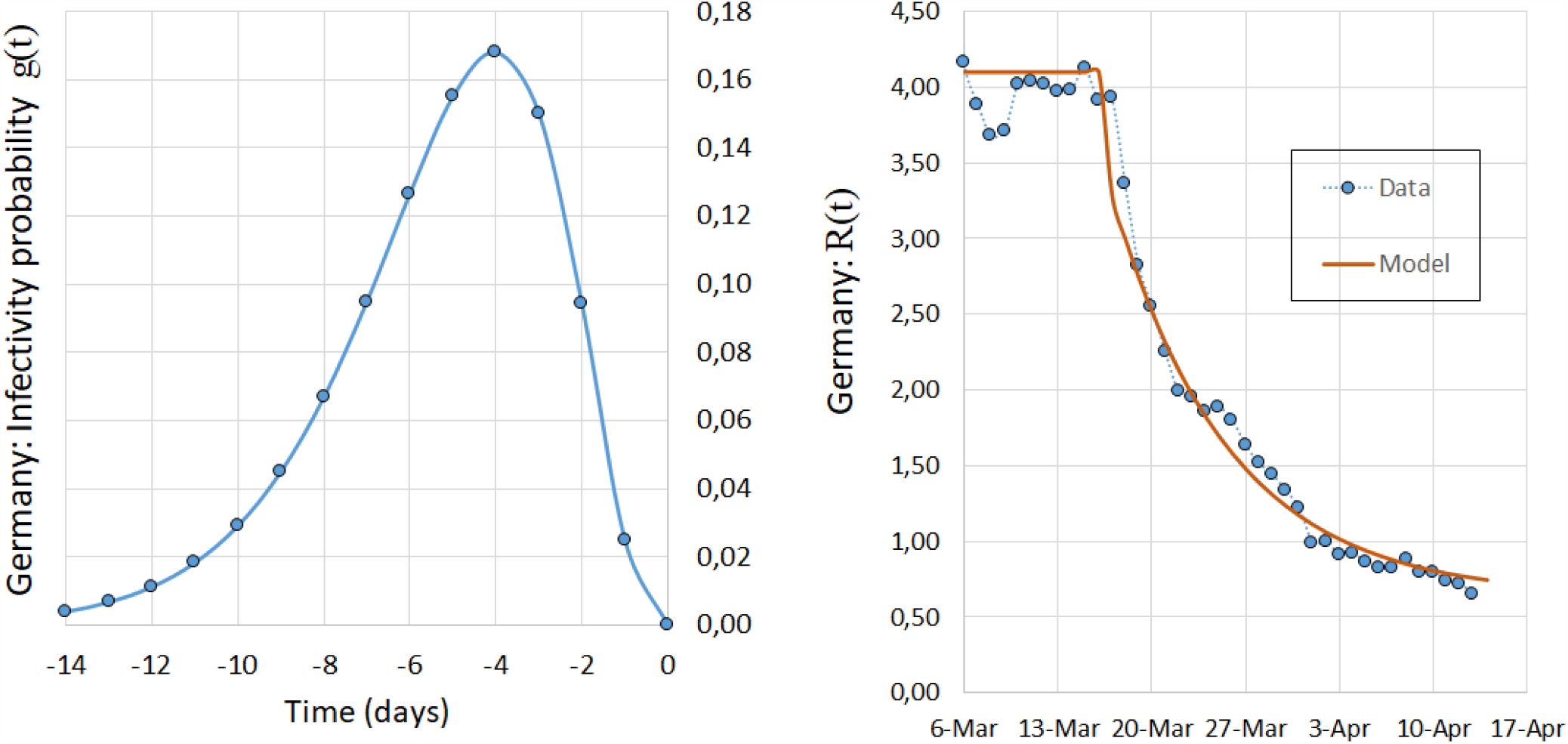
*Left: Γ -distributed weights used to calculate* 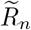 *with p* = 4 *and b* = 0.75 *(see eq*. (4)*). Right: Effective reproduction number R*(*t*) *obtained from data* 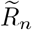 *(blue line) and the fitted curve (see eq*. (2)*) (orange line) used to model the epidemic in Germany. R*_0_ = 4.10, *α* = 0.12, *R*_*∞*_ = 0.64.

The effective reproduction number *R*(*t*) is shown for the time period from March, 3 to April, 14. The most restrictive measure in Germany was the school closing on March, 14. Note that some days before the 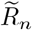 show a short plateau at *R*_0_ *≈* 4.0, which can be interpreted as the initial basic reproduction number *R*_0_ of COVID-19 in Germany. After three weeks, 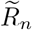 tends to the asymptotic value of about *R*_*∞*_ *≈* 0.6. We modelled this behaviour from day March 3, 2020 according to eq. (1), the parameters being: *R*_0_ = 4.10, *α* = 0.12, *R*_*∞*_ = 0.64. If we compare this to the analysis of Italy, we note that

1. The initial effective reproduction number *R*_0_ was higher in Germany;
2. Its final value *R*_*∞*_ is higher in Italy;
3. The rate parameter *α* is in both countries approximately the same.

A tentative interpretation can be the following: The initial reproduction number in Germany was higher because at the beginning of the epidemic the disease spread mainly among young people coming from skiing resorts in the Austrian Alps. If we assume that social contacts among young and sporty people are more frequent, this could be an explanation.

Although the measures taken by Italian politicians were more restrictive than in Germany, the effective reproduction number *R*_*∞*_ at mid April was 30% higher in Italy, leading to a much slower decay of daily new positive cases *ΔP* (*t*).

#### The COVID-19 cases in Germany

The number of daily positive diagnosed cases are shown on the left of Fig. 7. Original data are shown as a 7-day averages (blue dots). The model function (orange line) is capable of reproducing the course of the epidemic correctly but the peak value at March, 30 is underestimated. On the right panel the model function for the cumulative number of positive diagnosed cases *P* (*t*) reproduces almost exactly the time course of the data 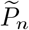.

**Fig 7:**
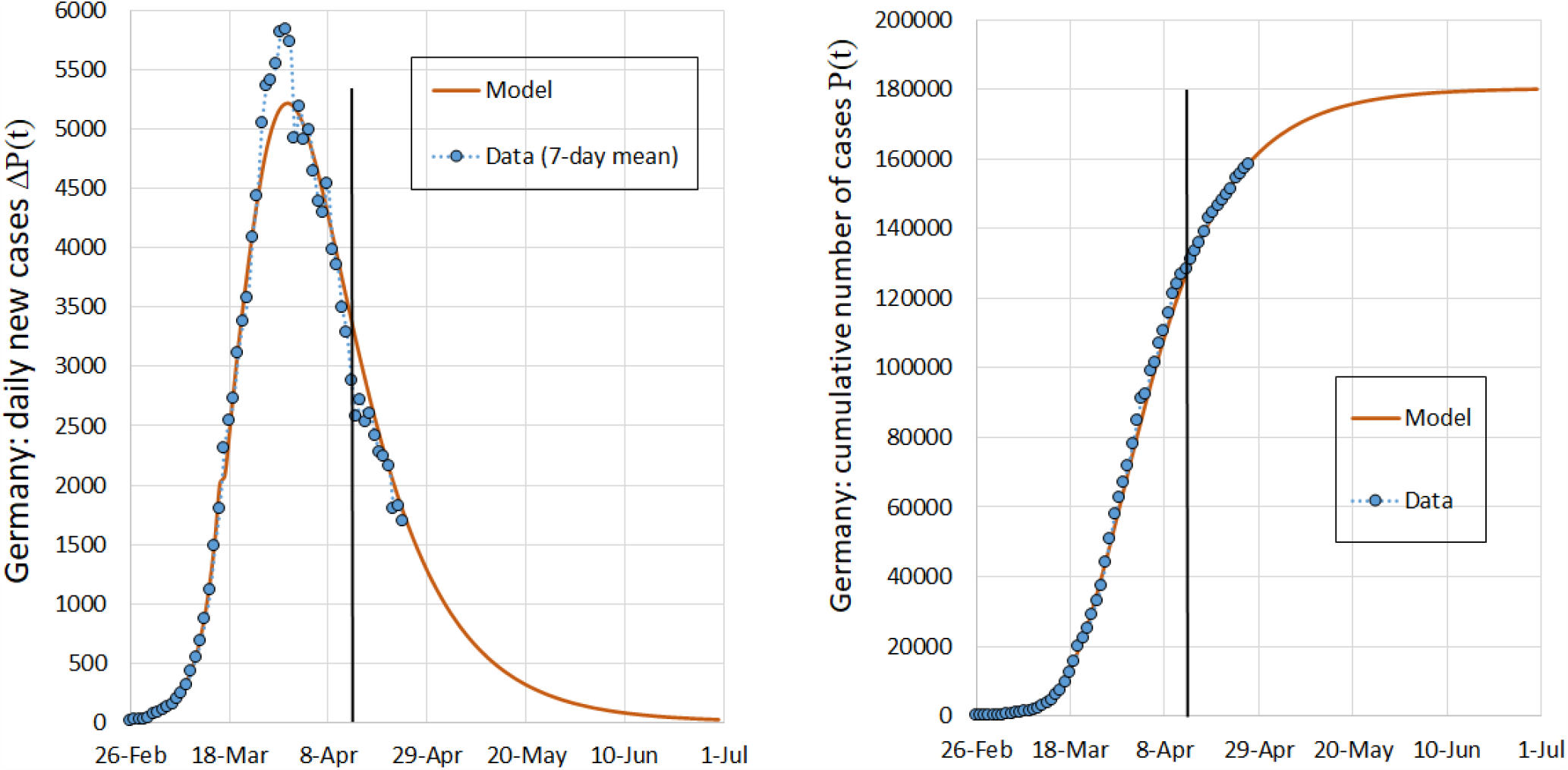
*Daily positive diagnosed cases ΔP* (*t*) *(left) and number of cumulative positive diagnosed cases P* (*t*) *(right) from February 26 to July 1, 2020 in Germany. Only data before the vertical black line, i*.*e*., *before April 13, have been used to tune the model*.

#### Fatalities in Germany

On the left of Fig. 8 we see the Gaussian weights we used to obtain the case fatality rates 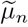 shown on the right panel. The integration time is *N*_*r*_ = 18 days. Note a time shift of the Gaussian by *t*_shift_ = 10 days putting a maximum weight on patients that have been diagnosed positive 10 days earlier, which corresponds to the average time in hospital before deceasing given by the Robert-Koch Institut [4].

**Fig 8:**
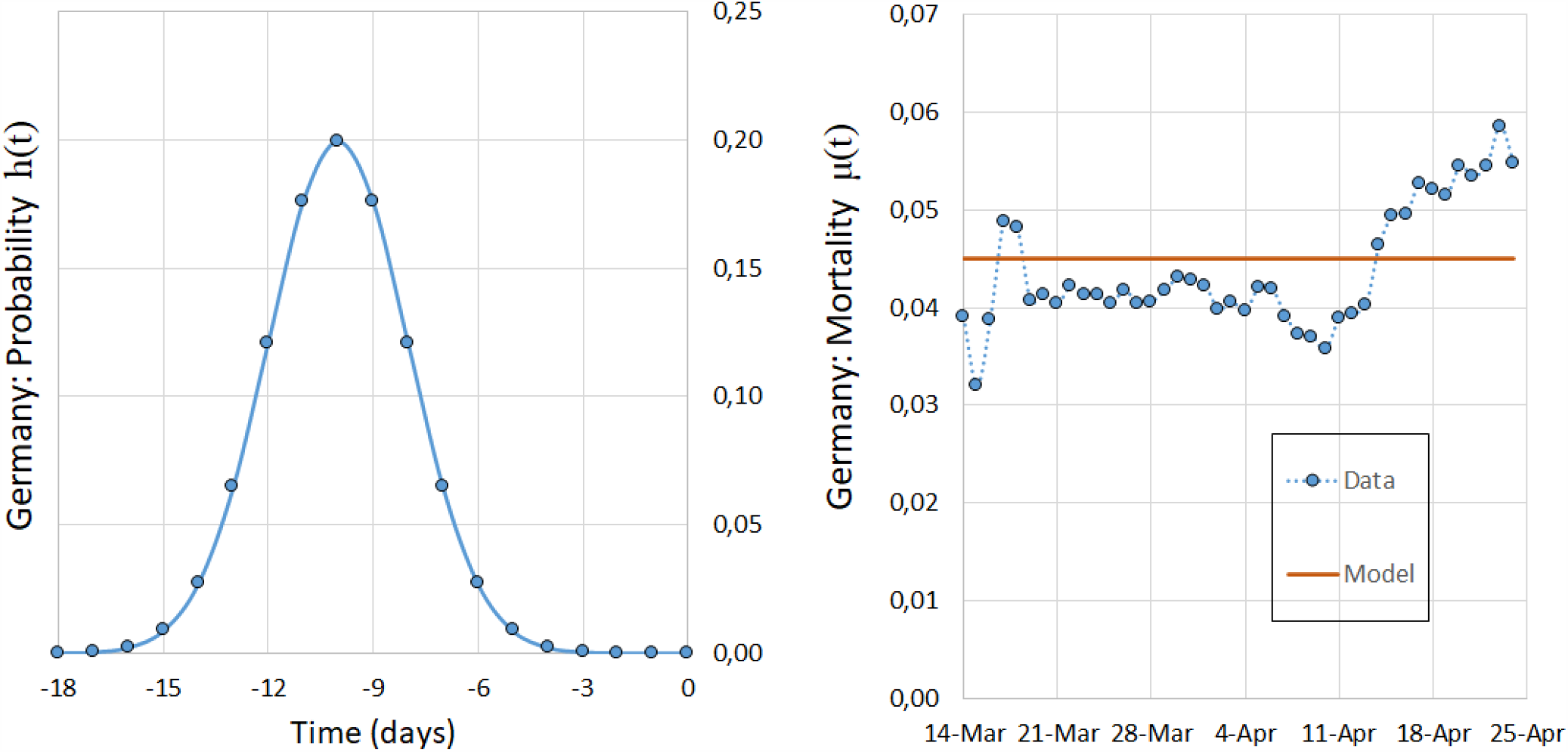
*Left: Gaussian weights h*_*i*_ *used to calculate* 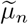 *with σ* = 2 *and t*_shift_ = 10. *Right: case fatality rate µ*(*t*) *obtained from data (blue points) and the fitted curve (orange line) used to model the epidemic in Germany*.

The number of daily and cumulative fatalities for Germany are shown in Fig. 9. The course of fatalities was not well represented due to a sudden rise in case fatality rate about April, 15. This effect is also visible on the right panel of Fig. 8, where there is a sudden increase in the mortality rate. For simplicity we assumed a constant value *µ*(*t*) = 0.045 representing the mean value between the two phases.

**Fig 9:**
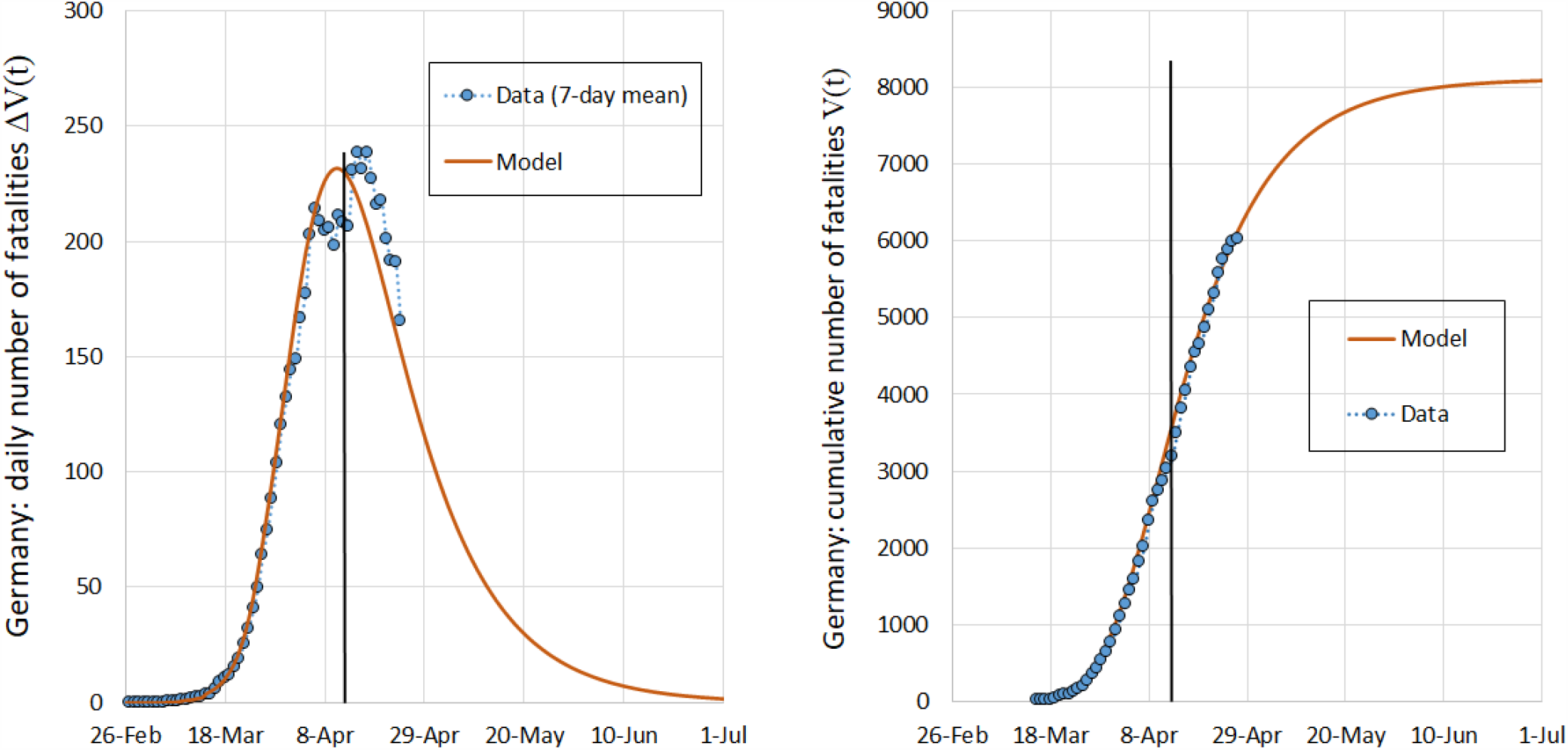
*Daily new fatalities ΔV* (*t*) *(left) and cumulative number of fatalities V* (*t*) *(right) from February 26 to July 1, 2020 in Germany. Only data before the vertical black line, i*.*e*., *before April 13, have been used to tune the model*.

#### Other Variables

Once the function describing the cumulative positive diagnosed cases *P* (*t*) is known, other variables can be derived, such as the number of:

− cured individuals,
− hospital beds needed,
− intensive care units (ICU’s) needed,
− active and closed cases,

and so forth. Forecasting these variable is made with an analogous weighted-integral-approach as for the fatalities, see Sect. 2.3. In Fig. 10 we show an example for the number of cumulative cured and active cases for Germany. Note that the same diagram for Italy (not shown) was difficult to obtain because the data of cured cases appeared not to be reliable. In fact, cured individuals were registered with a very long time delay (as to date of writing, cured cases made up only 65% of all closed cases).

**Fig 10:**
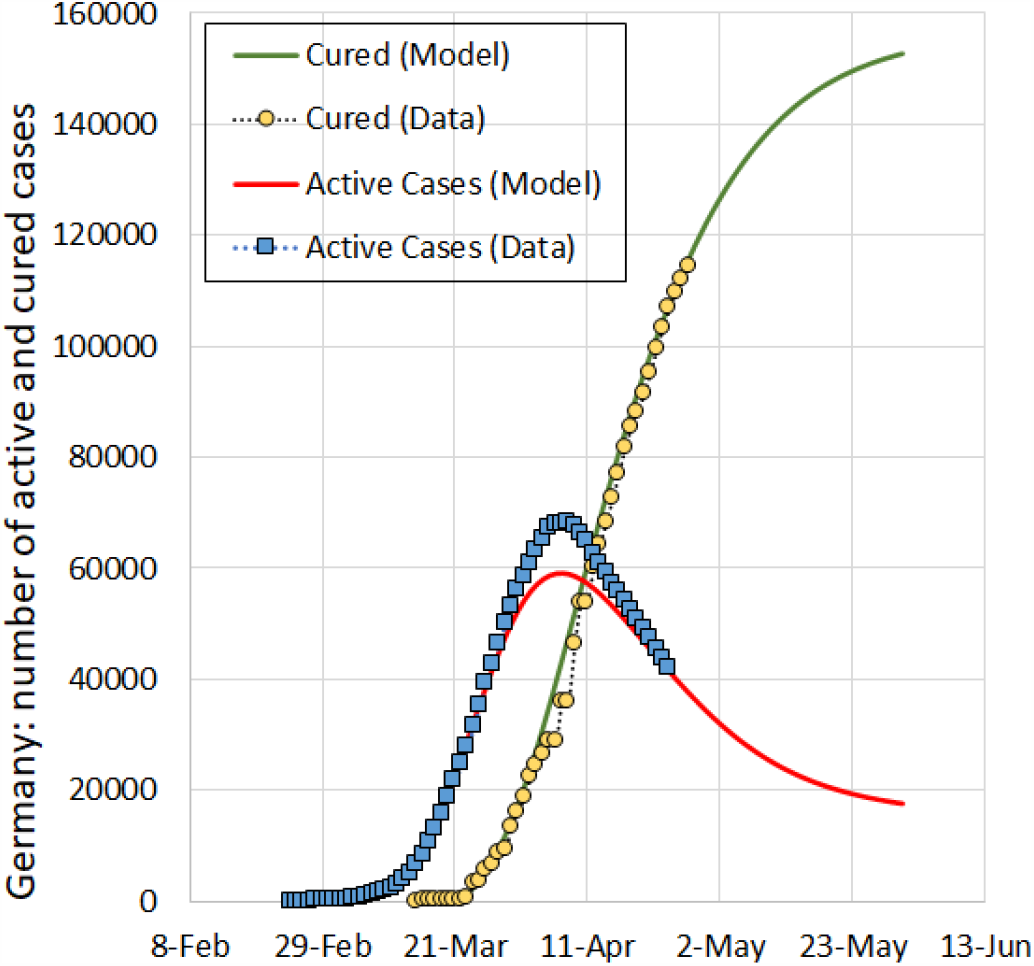
*Number of cumulative cured cases C*_*n*_ *from data (yellow circles) and the model simulation C*(*t*) *(green line), along with the number of active cases from data I*_*n*_ *(blue squares) and the model simulation I*(*t*) *(red line) from February 26 to June 5, 2020 in Germany*

## 4 Other Countries

We have applied our model to the COVID-19 data made available for other countries by the European Centre for Disease Prevention and Control [27]. The corresponding graphs are available on a web platform [1]. Here, we simply summarise the fitting parameters in table 1. The date of fitting was April 28, 2020.

**Table 1:** Fitting parameters for the effective reproduction number *R*(*t*).

| Country | $R_0$ | $\alpha$ | $R_\infty$ |
| --- | --- | --- | --- |
| Austria | 3.4 | 0.065 | 0.50 |
| Belgium | 3.2 | 0.045 | 0.64 |
| Brazil | 3.6 | 0.075 | 1.04 |
| France | 3.4 | 0.046 | 0.52 |
| Germany | 4.1 | 0.12 | 0.64 |
| Iran | 4.7 | 0.12 | 0.93 |
| Italy | 2.8 | 0.12 | 0.75 |
| Russia | 3.0 | 0.018 | 0.50 |
| South Korea | 2.3 | 0.34 | 0.76 |
| Spain | 2.9 | 0.074 | 0.63 |
| Sweden | 3.4 | 0.18 | 1.14 |
| Turkey | 3.9 | 0.10 | 0.84 |
| UK | 3.2 | 0.032 | 0.50 |
| USA | 2.8 | 0.12 | 0.91 |

**Table 2:** Model reaction to a 5%-perturbation of the fitting parameters.

| | $\delta P$ (2 weeks) | $\delta P$ (4 weeks) | $\delta P$ (6 weeks) | $\delta P$ (8 weeks) |
| --- | --- | --- | --- | --- |
| $\delta\alpha = +5\%$ | 1% | 15% | 21% | 22% |
| $\delta R_0 = +5\%$ | 20% | 21% | 21% | 21% |
| $\delta R_\infty = +5\%$ | 0% | 4% | 7% | 9% |

**Table 3:** Model reaction to a 10%-perturbation of the fitting parameters.

| | $\delta P$ (2 weeks) | $\delta P$ (4 weeks) | $\delta P$ (6 weeks) | $\delta P$ (8 weeks) |
| --- | --- | --- | --- | --- |
| $\delta\alpha = +10\%$ | 2% | 26% | 35% | 38% |
| $\delta R_0 = +10\%$ | 43% | 46% | 47% | 47% |
| $\delta R_\infty = +10\%$ | 0% | 8% | 16% | 21% |

The following facts can be observed:

− The lowest final reproduction number is 0.5.
− Some countries, such as Brazil and Sweden, still had *R*(*t*) *>* 1.
− The highest slope can be seen with South Korea, that has quickly introduced severe measures.

## 5 Stability of the model

Concerning the numerical error propagation for our model, we expect a high stability due to its integral formulation. This is important when applying the model to noisy time series data because the parameters that govern the overall dynamic are not obtained from a single data point but from a weighted sum of data points, see eq. (1). We cannot make a theoretical stability analysis and therefore we limit ourselves to show empirically how *P* (*t*) reacts to small changes in the curve fitting parameters. To this purpose, we individually disturb each of the parameters *R*_0_, *α* and *R*_*∞*_ associated with the fitting curve *R*(*t*) in (2) by 5% and observe the resulting change *δP* in *P* (*t*) after 2, 4, 6 and 8 weeks. The results are given in the following table:

Model deviations remain reasonably limited for mid-term forecasts up to two months. If we double the perturbation to 10% we observe the following changes of the cumulative number of diagnosed positive cases *P* (*t*):

It can be noted that doubling the perturbation roughly leads to the doubling of the deviation of the function. Thus, the model can be seen as numerically robust. The same results are expected for secondary variables, such as fatalities because they depend linearly on *P* (*t*).

## Conclusions

The main advantage of our approach is the simplicity of its formulation, the precision with which the real course of significant variables can be reproduced and the effectiveness to make mid-term forecasts.

We were able to show that our model has similarities to classic compartmental models, in particular:

− The variable *R*(*t*) can be interpreted as an effective reproduction number.
− The limited integration interval in the deterministic equation models the removal process.
− The *Gamma* −distributed weights account for infectiousness within a latency period, corresponding to the exposed state of the SEIR model.

Since our model consists of only one deterministic equation it is simpler compared to most approaches but is nevertheless able to capture the time course of the epidemic. In addition, the integral formulation leads to a good numerical robustness.

The model contains six parameters: three parameters related to infectiousness, *N*_*r*_, *b* and *p*, can be set from biological/clinical data typical of the disease; three free parameters, *α, R*_0_ and *R*_*∞*_ that can be obtained by the fitting procedure of the set of sample data 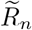. In the case of extinction of the epidemic outbreak, the parameter *R*_*∞*_ is expected asymptotically to vanish. A non-null value of *R*_*∞*_ indicates that the epidemics remain “latent”, with a relative small number of daily positive diagnosed cases for a very long time. This is indeed what seems to happen in various countries where the primary outbreak has been contained by adopting (even severe) social restrictive measures. Hence, deviations from a reliable forecasted value of *R*_*∞*_ could be intepreted as a warning signal of oncoming second epidemic wave [28].

We applied our model to many countries with a special focus on the data of Italy and Germany. After extracting the three fitting parameters we have been able to model the course of the epidemic in both countries rather well. We found it interesting how the parameters *R*_0_ and *R*_*∞*_ differ between the two countries - inviting us to interpret them in terms of effectiveness of measures, social organisation (in Italy elderly vulnerable people are more likely found to live with the younger part of the family) as well as the organisation of the health system.

We set up the hypothesis that three parameters suffice to model the epidemic from the outbreak, over the period of social distancing measures until the end - under the assumption that the measures remain effective with respect to infections till the end, *i*.*e*., zero new infections. It remains to be shown that this hypothesis remains valid for longer periods of time, especially when mitigation measures are loosened.

We hope that our approach facilitates forecasting of ongoing epidemics for long term periods, providing early warnings of further epidemic waves. Updating the model and re-tuning is very easy and we collect the results on the website [1].

## Data Availability

The data we used are official data made available by the John-Hopkins-University and the web site worldometers.info

https://coronavirus.jhu.edu/map.html

https://www.worldometers.info/coronavirus/

## Acknowledgement

We wish to thank Prof. Klaus Dietz for a very useful and illuminating discussion.

## Notes

### Competing Interest Statement

The authors have declared no competing interest.

### Funding Statement

No funding was received for this research

### Author Declarations

exemption for the research described

## References

1. http://www.roehlnet.de/corona/countries-all

2. I. Ciufolini, A. Paolozzi, A Mathematical prediction of the time evolution of the Covid-19 pandemic in some countries of the European Union using Monte Carlo simulations, Eur Phys J Plus. 135(4) 355 (2020)

3. http://web.math.unifi.it/users/brugnano/covid19/

4. M. an der Heiden, U. Buchholz, Modellierung von Beispielszenarien der SARS-CoV-2-Epidemie 2020 in Deutschland, Report, Robert Koch-Institut, Germany (2020), https://edoc.rki.de/bitstream/handle/176904/6547/Modellierung SARS-CoV-2.pdf?sequence=1

5. G. Giordano, F. Blanchini, R. Bruno, P. Colaneri, A. Filippo, A. Matteo, A SIDARTHE Model of COVID-19 Epidemic in Italy, 2003.09861 [q-bio.PE] (2020)

6. S. Uhlig, K. Nichani, C. Uhlig, K. Simon, Modeling projections for COVID-19 pandemic by combining epidemiological, statistical, and neural network approaches, medRxiv. https://www.medrxiv.org/content/early/2020/04/22/2020.04.17.20059535 (2020)

7. J. Wallinga, P. Teunis, Different epidemic curves for severe acute respiratory syndrome reveal similar impacts of control measures, Am. J. Epidemiol. 160(6), 509–516 (2004)

8. S. Cauchemez, P.Y. Boelle, G. Thomas, Estimating in real time the efficacy of measures to control emerging communicable diseases, Am. J. Epidemiol. 164(6), 591–597 (2006)

9. A. Cori, N.M Ferguson, C. Fraser, S. Cauchemez,caucheme A new framework and software to estimate time-varying reproduction numbers during epidemics Am. J. Epidemiol. 178(9), 1505–1512 (2013)

10. L. Dell’Anna, Solvable delay model for epidemic spreading: the case of Covid-19 in Italy, 2003.13571 [q-bio.PE] (2020)

11. A. Elazzouzi, A. Lamrani Alaoui, M. Tilioua, A. Tridane, Global stability analysis for a generalized delayed SIR model with vaccination and treatment, Adv. Differ. Eq. 2019, 532 (2019)

12. N. Shao, M. Zhong, Y. Yan, H. Pan, J. Cheng, W. Chen, Dynamic models for Coronavirus Disease 2019 and data analysis, Math. Methods Appl. Sci. 43(7), 4943–4949, (2020)

13. C. Heneghan, J. Brassey, T. Jefferson, COVID-19: What proportion are asymptomatic? Center for Evidence-Based Medicine, University of Oxford, https://www.cebm.net/covid-19/covid-19-what-proportion-are-asymptomatic/ (2020)

14. J. Wallinga, M. Lipsitch, How generation intervals shape the relationship between growth rates and reproductive numbers, Proc. Biol. Sci. 274, 599–604 (2007)

15. S. Flaxman, S. Mishra, A. Gandy, Estimating the number of infections and the impact of nonpharmaceutical interventions on COVID-19 in 11 European countries, https://www.imperial.ac.uk/mrc-global-infectious-disease-analysis/covid-19/report-13-europe-npi-impact/

16. H. Andersson, T. Britton, Stochastic epidemics in dynamic populations: quasi-stationarity and extinction, J. Math. Biol. 41(6), 559–580 (2000)

17. C.M. Hernández-Suarez, C. Castillo-Chavez, O.M. López, K. Hernández-Cuevas, An application of queuing theory to SIS and SEIS epidemic models, Math. Biosci. Eng. 7(4), 809–823 (2010)

18. W. Feller, On the integral equation of renewal theory, Ann. Math. Stat. 12, 243–267 (1941)

19. W.O. Kermack, A.G. McKendrick, A contribution to the mathematical theory of epidemics, Proc. Royal Soc. London 115, 700–721 (1927)

20. H.W. Hethcote, The mathematics of infectious diseases, SIAM Rev. 42(4), 599–653 (2000)

21. G. H. Li, Y. X. Zhang, Dynamic behaviors of a modified SIR model in epidemic diseases using nonlinear incidence and recovery rates, PLoS One, 12(4), e0175789 (2017)

22. P. van den Driessche, Some epidemiological models with delays, In: Differential Equations and Applications to Biology and to Industry, 507–520 (World Sci. Publishing, River Edge, NJ, 1996)

23. J. Arino, P. van den Driessche, Time Delays in Epidemic Models. In: Arino O., Hbid M., Dads E.A. (eds) Delay Differential Equations and Applications, NATO Science Series (II. Mathematics, Physics and Chemistry), vol 205, (Springer, Dordrecht,2006)

24. O.S. Miettinen, Epidemiological research: terms and concepts, (Springer, Dordrecht Heidelberg London New York, 2011)

25. https://coronavirus.jhu.edu/map.html. Accessed on 28-04-2020

26. https://www.worldometers.info/coronavirus. Accessed on 28-04-2020

27. https://data.europa.eu/euodp/de/data/dataset/covid-19-coronavirus-data. Accessed on 28-04-2020

28. L. López, X. Rodó, The end of social confinement and COVID-19 re-emergence risk, Nat. Hum. Behav. https://doi.org/10.1038/s41562-020-0908-8 (2020).

